# A *ONECUT1* regulatory, non-coding region in pancreatic development and diabetes

**DOI:** 10.1101/2024.07.23.24310605

**Authors:** Sarah Merz, Valérie Senée, Anne Philippi, Franz Oswald, Mina Shaigan, Marita Führer, Cosima Drewes, Chantal Allgöwer, Rupert Öllinger, Martin Heni, Anne Boland, Jean-François Deleuze, Franziska Birkhofer, Eduardo G Gusmao, Martin Wagner, Meike Hohwieler, Markus Breunig, Roland Rad, Reiner Siebert, David Alexander Christian Messerer, Ivan G. Costa, Fernando Alvarez, Cécile Julier, Sandra Heller, Alexander Kleger

**Affiliations:** Institute of Molecular Oncology and Stem Cell Biology, Ulm University Hospital, Ulm, Germany; Université de Paris, Institut Cochin, INSERM U1016, CNRS UMR-8104, Paris, France; Department of Internal Medicine 1, Ulm University Hospital, Ulm, Germany; Institute for Computational Genomics, RWTH Aachen University Medical School, Aachen, Germany; Institute for Clinical Transfusion Medicine and Immunogenetics, German Red Cross Blood Transfusion Service Baden-Württemberg-Hessen and University Hospital Ulm, Ulm, Germany; Institute of Human Genetics, Ulm University & Ulm University Medical center, Ulm, Germany; Institute of Molecular Oncology and Functional Genomics, Center for Translational Cancer Research and Department of Medicine II, School of Medicine, Technical University of Munich, Munich, Germany; Division of Endocrinology and Diabetology, Department of Internal Medicine 1, Ulm University Hospital, Ulm, Germany; Institute for Clinical Chemistry and Pathobiochemistry, Department for Diagnostic Laboratory Medicine, University Hospital Tübingen, Germany; Université Paris-Saclay, CEA, Centre National de Recherche en Génomique Humaine (CNRGH), Evry, France; Centre of Informatics, Federal University of Pernambuco, Recife, Brazil; Institute for Transfusion Medicine, University Hospital Ulm, Ulm, Germany; Division of Gastroenterology, Hepatology & Nutrition, CHU Sainte-Justine, University of Montreal, Canada; Division of Interdisciplinary Pancreatology, Department of Internal Medicine 1, Ulm University Hospital, Ulm, Germany; Core Facility Organoids, Ulm University, Germany

## Abstract

In a patient with permanent neonatal syndromic diabetes clinically similar to cases with *ONECUT1* biallelic mutations, we identified a disease-causing deletion located upstream of *ONECUT1*. Through genetic, genomic and functional studies we identified a crucial regulatory region acting as an enhancer of *ONECUT1* specifically during pancreatic development. This enhancer region contains a low-frequency variant showing strong association with type 2 diabetes and other glycemic traits, thus extending the contribution of this region to common forms of diabetes. Clinical relevance is provided by experimentally tailored therapy options for patients carrying *ONECUT1* coding or regulatory mutations.

## Introduction

Diabetes mellitus has become a global major health burden. The disease is characterized by considerable clinical and etiological diversity. However, the current diabetes classification does not fully capture the disease’s heterogeneity^1^. Besides common forms of diabetes (T1D and T2D) having a multifactorial etiology with a polygenic background^2^, a subset of diabetes cases may result from single gene mutations. So far, numerous genes whose mutations cause monogenic diabetes forms have been identified and functionally characterized, revealing various mechanisms that disrupt β-cell integrity^3^. Interestingly, common variants in several of these monogenic diabetes genes have also been associated with polygenic predisposition to diabetes. This genetic overlap in common polygenic and rare monogenic diabetes forms suggests that disease mechanisms and biological pathways are partly shared between different forms of diabetes^4–12^. However, more than 90% of single nucleotide polymorphisms (SNPs) associated with diabetes identified through genome-wide association studies (GWAS) are situated outside coding regions of genes and can influence cellular function and the development of pancreatic islets^13–17^.Hence, dysfunctional gene regulatory elements (GRE), like promoters, enhancers, silencers, insulators, or transcribed regulatory sequences, such as long non-coding RNAs (lncRNA) could explain part of the missing heritability of type 2 diabetes^18^. Indeed, such SNPs display a notable frequency in pancreatic islet-enhancer clusters, highlighting the relevance of islet-specific gene regulation in the pathophysiology of T2D^13,19,20^. Nevertheless, comprehensive studies on regulatory elements driving metabolic disorders are relatively rare, but more such elements are being discovered^21–24^. Understanding mutations in genes and their regulatory regions causing severe monogenic diabetes can thus illuminate the path towards a more personalized diabetes medicine^19^.

Recently, we identified *ONECUT1* coding mutations as cause of a monogenic, recessively inherited syndromic form of diabetes with neonatal or very early onset. Additionally, rare coding variants of *ONECUT1* define a subgroup of patients with early-onset T2D and a favorable response to treatment. On the other hand, common regulatory variants near *ONECUT1* are associated with multifactorial T2D. Functional studies based on human pluripotent stem cell differentiations demonstrate that ONECUT1 plays a crucial role in pancreatic progenitor (PP) formation and endocrine development, affecting transcriptional regulation and epigenetic processes across various forms of diabetes^7,12^.

Here, we studied a child with neonatal syndromic diabetes and a family history of T2D whose clinical presentation was very similar to cases with biallelic *ONECUT1* truncating mutations. Genetic and genomic studies of the patient and his family followed by functional assays in genome-edited human embryonic stem cells (hESC) forming stem cell-derived islets (SC-islets) identified a novel *ONECUT1* regulatory element, characterized the disease-causing mechanism and experimentally underpinned clinical relevance of our findings.

## Results and Discussion

We studied a patient born to non-consanguineous parents with neonatal syndromic diabetes, associated with intrauterine growth retardation and pancreatic and hepatic manifestations (more details in *Methods*). Weight at birth was <5th percentile and height <1st percentile. Diabetes was diagnosed at 0-5 years, requiring insulin thereafter. He also had exocrine pancreas insufficiency (very low fecal chymotrypsin), treated by pancreatic enzymes. He had failure to thrive and growth retardation (height and weight <1st percentile). Ultrasound showed a very hypoplastic pancreas and the absence of gall bladder, similar to neonatal diabetes patients carrying biallelic *ONECUT1* mutations^7,25^ (e.g. **Fig.1a,b**). Using Sanger sequencing of *ONECUT1* exons in this patient and his parents, we identified a heterozygous frameshift mutation in *ONECUT1* (ONECUT1-p.A206Tfs58X) in the patient. This mutation results in a truncated protein lacking the CUT and HOX domains and was inherited from his mother (**Fig.1c**). The family history of adult-onset diabetes (grandparent) suggests the involvement of this protein-truncating variant (PTV) in adult-onset diabetes in line with our recent findings^7,12,25^. Considering the severe neonatal presentation of the patient (comparable with biallelic *ONECUT1* mutation phenotype), we performed high-density SNP array genotyping, whole genome sequencing (WGS) and confirmatory Sanger sequencing in the patient and his parents and identified a deletion of 108 kb (chr15.hg19:g.53694760_53802787del) located 612 kb upstream of *ONECUT1*, transmitted by the father (**Fig.1c,d, Methods)**. This non-coding region contains a lncRNA gene (AC066614.1; further labeled lncRNA-AC) as well as binding peaks of ONECUT1, FOXA1/2, GATA6, NKX6.1 and PDX1 (**Fig.1d**), supporting its putative role in *ONECUT1* regulation.

**Figure 1:**
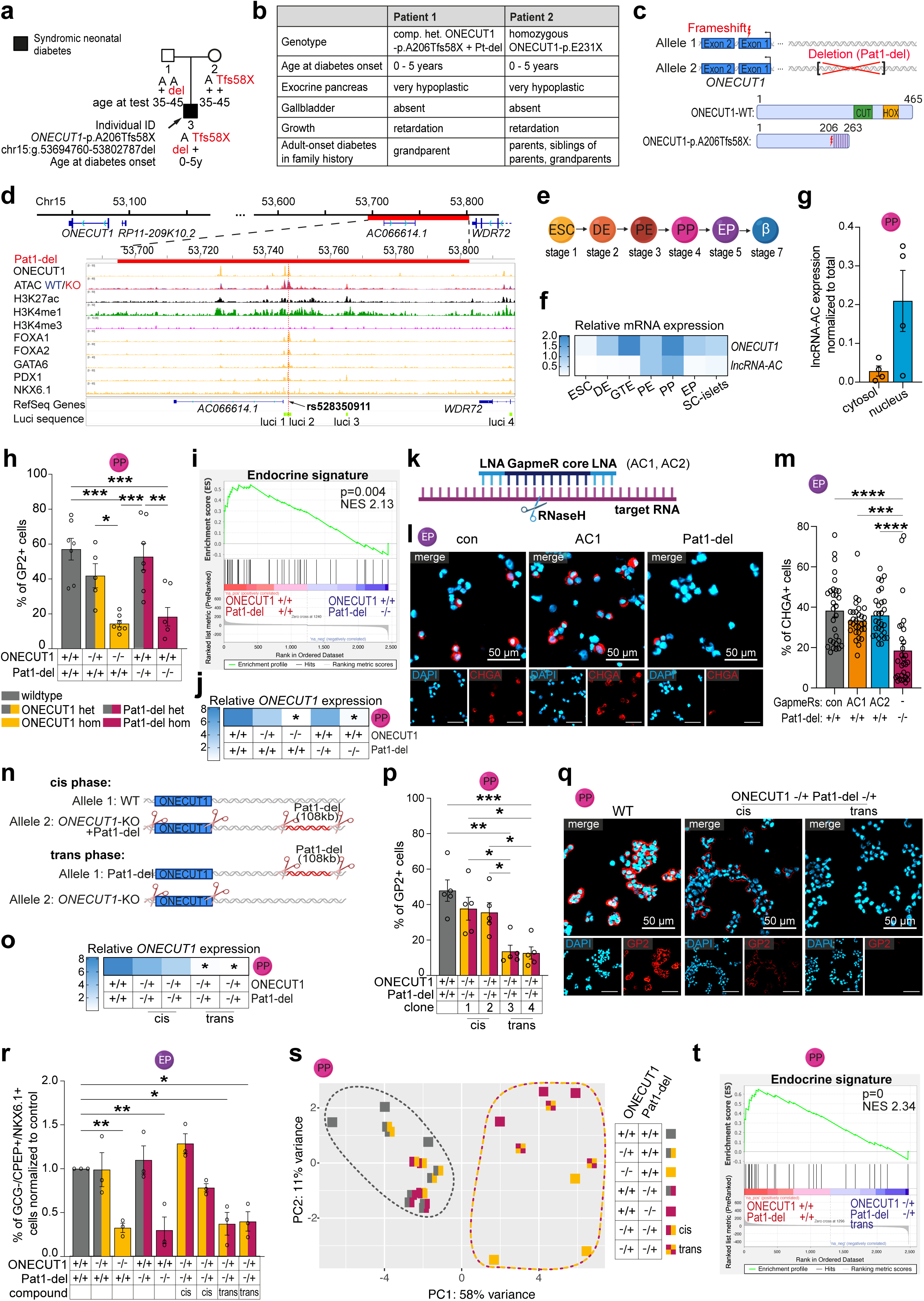
A *ONECUT1* regulatory non-coding region during pancreatic development and endocrine specification. (**a**) Pedigree describing a neonatal syndromic diabetes patient 1 and the family. (**b**) The clinical presentation of patient 1 resembles previously reported cases with biallelic *ONECUT1* mutations, here compared with a neonatal diabetes patient (patient 2) carrying a homozygous *ONECUT1* truncating mutation^7^. (**c**) Schematic genotype of the patient and consequenses of the missense mutation on the ONECUT1 coding region. (**d**) IGV plot of the region deleted in patient 1 (Pat1-del) with corresponding ChipSeq peaks of pancreatic TFs, histone marks and ATAC peaks from PP stage of WT and *ONECUT1* deleted hESCs^7^. The annotated lncRNA is marked (lncRNA-AC; AC066614.1). (**e**) Schematic stepwise pancreatic differentiation of hESCs to definitve endoderm (DE), gut tube endoderm (GTE), pancreatic endoderm (PE), pancreatic progenitors (PP), and endocrine progenitor (EP) cells and stem cell-derived islets (SC-islets). (**f**) qPCR of lncRNA-AC and *ONECUT1* during differentiation (n=3-4); normalized to PE. (**g**) Subcellular localization of lncRNA-AC determined by qPCRs (n=4). (**h**) Flow cytometry (FC) of the PP marker GP2 across indicated genotypes (n=5-7). (**i**) GSEA shows loss of an endocrine lineage gene set^31^ in Pat1-del PPs. (**j**) Relative *ONECUT1* expression in WT, *ONECUT1*- and Pat1-del cells at PP stage (n=3-4) based on qPCR. (**k**) Schematic outlining the GapmeR-experimental design. (**l,m**) GapmeR-based lncRNA-AC knockdown during differentiation and subsequent illustration of chromogranin A (CHGA)-positive cells via immunofluorescence (IF) and respective quantification. (**n**) Schematic genotype upon CRISPR-mediated deletion of the Pat1-del region in heterozygous ONECUT1-del hESCs. (**o**) *ONECUT1* expression was determined in cis and trans clones at PP stage (n=3-4). (**p,q**) GP2 expression determined by FC and IF at PP stage, respectively (n=5). (**r**) Amount of immature β-cells at EP measured via FC for NKX6.1, CPEP, GCG (n=3). (**s**) Principal component analysis (PCA) of whole transcriptomes. (**t**) GSEA shows loss of an endocrine lineage gene set^31^ in trans-deleted PPs.

Thus, we first hypothesized that the lncRNA-AC, which is mostly localized in the nucleus and also resembles the *ONECUT1* expression pattern during pancreatic differentiation (**Fig.1e-g**), mechanistically contributes to the glycemic phenotype of the patient. To explore this in greater detail, we deleted the patient-specific 108 kb region (referred to as Pat1-del, heterozygous and homozygous) in hESC and assessed their subsequent differentiation into PPs (**Fig.1e**). While the earlier stages of pancreatic development remained unaltered (**ExDataFig.1a**), the PP stage was strongly impaired in both homozygous Pat1-del or *ONECUT1* knockouts, as shown by reduced expression of PP markers (GP2, NKX6.1, PDX1; **Fig.1h**;**ExDataFig.1b,c**) as well as loss of pancreatic endocrine lineage and functional endocrine genes upon RNA-sequencing (**Fig.1i**;**ExDataFig.1d,e**). Of note, the patient’s homozygous deletion (Pat1-del -/-), almost completely suppressed *ONECUT1* expression, indicating strong regulation of *ONECUT1* transcription through this genomic region (**Fig.1j; ExDataFig.1f,g**).

LncRNAs have already been described to operate in concert with transcription factors (TF) regulating the transcriptional network in pancreatic β-cells^26^. To investigate a possible role of the lncRNA-AC transcript as a regulator of *ONECUT1* transcription, we utilized a GapmeR-based approach to knock down lncRNA-AC (**Fig.1k;ExDataFig.1h,i**). The decrease in lncRNA-AC levels did not affect differentiation efficiency, suggesting no direct role of the lncRNA-AC transcript (**Fig.1l,m;ExDataFig.1j,k,l**). Instead, we hypothesized that an enhancer activity, as indicated by pancreatic TF binding and histone modifications (**Fig.1d**), within the deleted non-coding region of the patient explains the drastic phenotype. Previous research indicates that neighboring genes of a lncRNA locus often interact through various mechanisms, such as RNA splice sites. This interaction can affect the function and evolution of these loci, regulating both coding and non-coding genes^27^.

To probe this hypothesis, we heterozygously deleted this 108 kb region in heterozygous *ONECUT1* KO hESCs (**Fig.1n**). Nanopore-sequencing confirmed either cis- (clone 1) or trans- (clone 3) orientation of the *ONECUT1* gene and its upstream regulatory element (**ExDataFig.2**). Among the four generated clones, two exhibited complete loss of *ONECUT1* expression as well as disruption of PP and endocrine progenitor (EP) formation, while the other two clones displayed a phenotype similar to single allelic deletions of either *ONECUT1* or the upstream located region (**Fig.1o-r;ExDataFig.3a-c**). Global transcriptome analysis similarly separated the genotypes (**Fig.1s,t;ExDataFig.3d**). Thus, we provide compelling genetic evidence that pinpoints a cis-regulatory element within a non-coding genomic region that acts as a transcriptional enhancer during pancreas development. Hence, the combined loss of *ONECUT1* and partial deletion of the enhancer region in a compound heterozygous status (trans phase) reproduces the severe phenotype observed in patient 1 which explains the strongly impaired PP formation capacity in ONECUT1 -/+ Pat1-del -/+ trans clones (**Fig.1a-c,q-t**).

We selected four regions (luci 1-4) from this non-coding element based on pancreatic TF-binding patterns (**Fig.1d**) and tested their minimal enhancer activity using luciferase assays. Interestingly, the fragment most enriched for ONECUT1 binding peaks displayed elevated reporter activity exclusively in the minus orientation (luci 2), which was even enhanced by ONECUT1 protein expression (**Fig.2a,b**). On the contrary, the plus-oriented fragment 2 (luci 2) exhibited repressive effects, as did fragments 1 and 3 (**Fig.2a,b**). Another fragment (luci 4), located further upstream of the 108 kb region deleted in patient 1, also comprised pancreatic TF peaks and showed similar induction patterns as luci 2 (**Fig.2a,b**) indicating at least two genomic sites with enhancer activity upstream of *ONECUT1*. Using nanopore-based 5mC detection, we identified differential hypomethylation from hESC to PP stage in part of the intron of *ONECUT1* (**ExDataFig.4**). Additionally, we observed differential hypomethylation in the identified enhancer region (including luci 2; **Fig.2c;ExDataFig.5**), which was associated with pancreatic TF binding (**Fig.1d)** and luciferase reporter activity (**Fig.2a**), indicating that hypermethylation keeps this enhancer inactive in non-pancreatic lineages.

**Figure 2:**
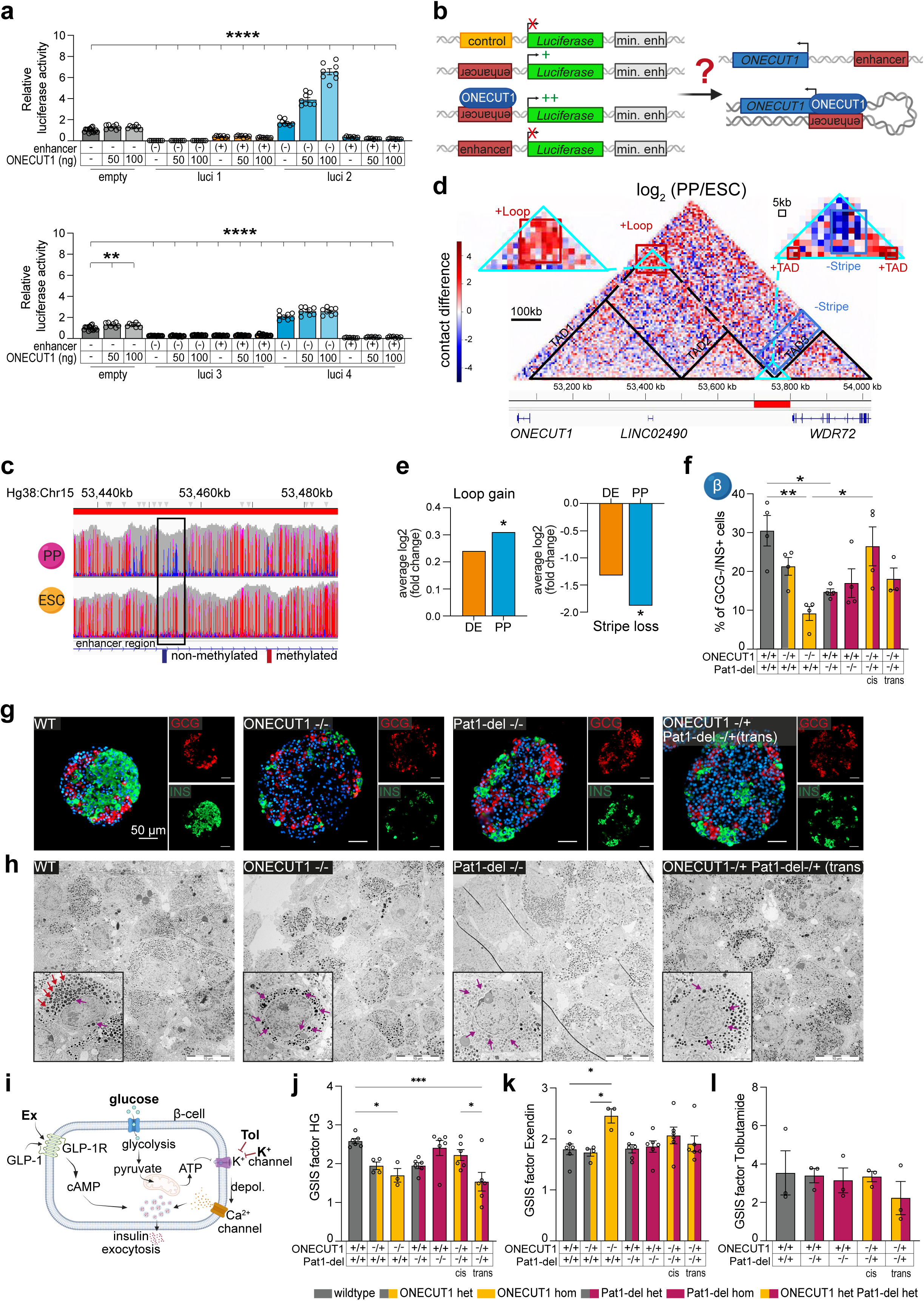
Functional characterization of the transcriptional enhancer. (**a**) Luciferase assays measuring enhancer activity of four distinct regions (luci 1-4) within and close to the 108 kb Pat1-del region (n=8). (**b**) Schematic of the + or – orientation of the putative enhancers and physical occupation by ONECUT1 (left). Proposed chromatin looping mechanism, bringing *ONECUT1* promoter and enhancer region into close proximity (right). (**c**) DNA methylation in hESC and PPs around the pancreatic enhancer determined via nanopore-sequencing. Boxed area marks the enhancer region (including luci 2). Color code gives amount of methylated cytosines (red) as compared to umethylated cytosines (blue). (**d**) Hi-C data analysis identifying structural changes between hESC and PP is depicted in a log2 fold-change map. Lost and gained structures are highlighted in blue and red, respectively. (**e**) The average log2 fold change of gained loop structure in its TAD (chr15:53065000_53765000) and for stripe loss in 100-kb region (chr15:53700000_53800000) are displayed. (**f**) Percentage of β-cells (INS+/GCG-) in SC-islets upon differentiation to stage 7 as measured by flow cytometry. (**g**) Representative immunofluorescence images of SC-islets showing GCG (red) and INS (green) positive cells in various genotypes. (**h**) Transmission electron microscopy overview images and representative images of single cells. Red arrow: mature electron-dense endocrine vesicles, located near the apical membrane; purple arrow: immature, less dense endocrine vesicles, located near the nucleus. (**i**) Schematic illustration of the insulin secretion machinery triggered by various stimuli, such as glucose, exendin-4 (Ex), KCl and tolbutamide (Tol). (**j,k,l**) Secreted insulin was measured in response to various stimuli, and depicted from left to right as follows: high glucose (HG; 20 mM) normalized to low glucose (LG, 1 mM); HG + 50 ng/ml Ex normalized to HG; LG + 400 µM Tol normalized to LG. n=3-6.

Assuming a differentially formed chromatin loop between the regulatory element and the *ONECUT1* promoter, we compared stage-specific Hi-C genomic sequencing data from hESC differentiated to the pancreatic lineage. Here, we identified progressive chromatin changes during pancreatic differentiation reaching significance at PP stage (**Fig.2d,e;ExDataFig.6a**). We identified a new chromatin loop forming from the extrusion process that combines topologically associating domaines (TAD)1 and TAD2, structurally linking the *ONECUT1* promoter with the regulatory element. We also observed the left boundary of TAD3 shifting to a new position within the adjacent *WDR72* locus, seen as a structural loss in the ESC stage and supported by relocated CTCF binding (**ExDataFig.6a,b**). Collectively, these findings suggest the establishment of a new chromatin loop in PP cells along the respective TAD2. Intriguingly, the boundary between TAD2 and TAD3 coincides with the regulatory element located within the patient 1-specific 108 kb region (**Fig.2d,e**). The emergence of this loop at the PP stage indicates stronger interaction between the *ONECUT1* promoter and the regulatory element, supporting the proposed mechanism of a distal cis-acting enhancer regulating *ONECUT1* expression during pancreatic development. In conclusion, deletion of this element impairs *ONECUT1* and represents a disease-causing allele to *ONECUT1* recessive neonatal diabetes.

To further explore the relevance of our finding to common T2D, we investigated public data from HugeAMP. We identified a low-frequency variant, rs528350911 (chr15.hg19:53747228-C-G, minor allele frequency [MAF] = 0.0035 in gnomAD, all populations) located within the putative enhancer region (precisely within luci 2) associated with increased risk of T2D and other glycemic traits, including random glucose (P=2.6×10^−8^, beta=0.081), and T2D adjusted for BMI (T2DadjBMI, 5.9×10^−7^, odds ratio [OR] =1.31) (**Tab.1**). This variant is located at the binding sites of several pancreas-specific TFs such as PDX1, NKX6.1, and ONECUT1 (**ExDataFig.6b**). Of note, the strength of diabetes association at this low-frequency variant is in the same order of magnitude as the strongest T2D-association signal over the whole genome (e.g. OR=1.34 for T2D and T2DAdjBMI at the TCF7L2 lead SNP rs7903146, compared to OR=1.38 and 1.31 respectively for rs528350911, HugeAMP), emphasizing the significance of this association. The detected association of this variant with T2D and other glycemic traits independent of BMI underscores our findings of a pancreatic phenotype that ultimately impairs insulin secretion and argues against a mechanism dependent on insulin sensitivity.

**Table 1:** Glycemic traits associated with a SNP (rs528350911) mapping to the identified enhancer region.

| Trait | P-value | OR | Beta | N |
| --- | --- | --- | --- | --- |
| Random glucose | 2.56E-08 |  | 0.081 | 400775 |
| T2D adjusted for BMI | 5.89E-07 | 1.31 |  | 175915 |
| HbA1c | 5.20E-05 |  | 0.051 | 438069 |
| T2D | 0.00057 | 1.38 |  | 183738 |

Association values and effect size (OR and beta) are shown for the rare variant of SNP rs528350911 (chr15:hg19:53747228-C-G), G allele (MAF=0.0035, gnomAD all populations). Data are extracted from HUGOAMP. OR: Odds ratio, N: size of the cohort (multi-ancestry).

Next, we investigated the mutant SC-islet upon further differentiation of the PPs (**ExDataFig.7a**) to the more mature endocrine stage 7^28^: Analysis of the SC-islet composition revealed reduced formation of β-cells in both heterozygous and homozygous Pat1-del hESCs as well as in the ONECUT1 -/+ Pat1 -/+ compound heterozygous clones, while the number of glucagon-, pancreatic polypeptide- or somatostatin-positive α-, γ- and δ-cells, remained unaffected (**Fig.2f,g;ExDataFig.7b-g**). The overall similar chromogranin A and vimentin levels per SC-islet excluded aberrant loss of lineage identity (**ExDataFig.8a,b**). Transmission electron microscopy revealed disturbed polarization, elevated levels of immature granules, and a decreased overall presence of electron-dense granules in the SC-islets carrying either a homozygous ONECUT1 KO or Pat1-del mutation (**Fig.2h**).

Next, we probed SC-islet functionality using glucose-stimulated insulin secretion assays (GSIS) and subsequent drug treatment (**Fig.2i**): GSIS demonstrated perturbed insulin secretion of the remaining β-cells in *ONECUT1* knockout and *ONECUT1* -/+ Pat1-del -/+ compound heterozygous lines (**Fig.2j;ExDataFig.8c,d**). It is noteworthy that single or compound-cis heterozygous clones exhibited also defective insulin secretion during GSIS (**Fig.2j**).

Finally, our goal was to evaluate potential rescue strategies that could improve impaired β-cell function in various genotypes affecting both the coding and regulatory, non-coding regions of the *ONECUT1* gene. Treatment of SC-islets with the GLP-1 receptor agonist exendin-4 stimulated insulin secretion in all genotypes, with homozygous ONECUT1 KO mutants exhibiting even better responses than WT counterparts (**Fig.2k;ExDataFig.8c,d**). Subsequent tests with the sulfonylurea tolbutamide yielded consistent responses across all genotypes (**Fig.2l;ExDataFig.8c,d**). Thus, despite the lower number of β-cells and the diminished glucose responsiveness of carriers of *ONECUT1* variants (coding or regulatory), their β-cells still respond to glucose-lowering drugs. This indicates the potential of GLP-1 receptor agonists or sulfonylureas as optimized treatment options for patients carrying mutations in *ONECUT1* coding and/or regulatory regions (**Fig.2k,l;ExDataFig.8c,d**).

In summary, our findings reveal the pathomechanism leading to neonatal diabetes in a patient with a compound heterozygous *ONECUT1* truncating variant and a deletion affecting key regulatory elements. This discovery highlights essential *ONECUT1* regulatory regions involved both in monogenic diabetes and in common T2D, emphasizing the transcription factor’s role in endocrine pancreas development. Variation in this locus may contribute to the specific T2D subtype characterized by low insulin secretion^1,29^.

During the preparation of this manuscript, the same enhancer region was identified and reported in an unbiased CRISPR screen, confirming and strenghtening our results presented within this study^30^.

## Data Availability

All data produced in the present study are available upon reasonable request to the authors.

## Acknowledgement

We thank Marc Nicolino and Kevin Perge from the Hospices Civils de Lyon and Cochin Institut for helpful discussions. We thank the patient and his family for participating in this study. We thank Katrin Jochmann, Ralf Köhntop, Katrin Köhn, Ulrike Mayr-Beyrle and Franziska Ott for their excellent technical support. Further we thank all the IMOS lab members for the valuable discussions. S.M. and S.H. are supported by DFG grant HE 7277/3-1 and the Medical Scientist Program at Ulm University awarded to S.H. This project was also funded by an ANR-DFG collaborative research project (grant no. ANR-18-CE92-0031, DFG KL 2544/5-1) to C.J and A.K, and by the European Foundation for the Study of Diabetes/JDRF/Novo Nordisk, the Assistance Publique-Hôpitaux de Paris Programme Hospitalier de Recherche Clinique (project DIAGENE) and France Génomique (project DIAPED) to C.J. The CNRGH sequencing platform was supported by the France Génomique National infrastructure, funded as part of the « Investissements d’Avenir » program managed by the Agence Nationale pour la Recherche (contract ANR-10-INBS-09).

CD is a scholar of the DFG-funded SFB1074 (project B10N).

## Author contribution

S.M. acquired data, analyzed and interpreted results, and drafted and revised the manuscript.

V.S. and A.P. contributed equally and performed the genetic studies, and analysed and interpreted the results, drafted the genetics section and revised the manuscript.

A.B. and J.F.D. generated the WGS and high density SNP array genotyping of the patient and his family on the CNRGH genomics platform.

F.A. identified, followed and clinically characterized the patient and his family.

F.B. generated Pat1-del knockout lines R.Ö. and R.R. performed RNA-seq

C.A. bioinformatically analyzed RNA-seq data and performed GSEA.

M.W. analyzed and interpreted TEM

F.O. conceptualized and performed luciferase assay

M.S., E.G.G. and I.G.C. bioinformatically analyzed and interpreted Hi-C data

M.F. and D.A.C.M. performed nanopore sequencing

C.D. and R.S. analyzed and interpreted ONT analyze phasing and methylation

M. Heni, M.B and M. Hohwieler interpreted data and revised the work.

C.J., S.H., A.K. (contributed equally) conceptualized and interpreted the work, and drafted and revised the manuscript.

All authors revised the manuscript and agreed for publication.

## Methods

### Patient with neonatal syndromic diabetes and his family

The index patient (patient 1) and his parents are described in the Results section, in **Fig.1a,b**, and below. The study was explained to the patient and his parents, who gave their written consent. Blood samples were collected from the patient and his parents and DNA extraction was performed using standard procedures. The study was approved by Sainte-Justine Hospital. Due to ethical restrictions, additional clinical informations on the patient are not described in detail. Details of interest can be provided by the lead author upon request.

#### Information on the parents and family

at the time of the patient’s study at 10-15 years of age, both parents were 30-40 years old and had no reported history of diabetes or treatment for the condition. However, their diabetes status could not be specifically investigated. One grandparent, aged 60-70, had T2D, though the age at diabetes onset was not known.

### *ONECUT1* mutation screening by Sanger sequencing

Sanger sequencing of ONECUT1 coding exons was performed in the patient and his two parents as described in ^7^.

### Genomic studies for patient’s deletion identification, with Sanger confirmation

High-density SNP genotyping and WGS were done at the Centre National de Recherche en Génomique Humaine (CNRGH), CEA.

DNA of the patient and his parents was genotyped using the Illumina Infinium Global Screening Array Multi-Disease v3.0 (GSA-MDv3.0) microarray. Genotyping call rate was greater than 99% (99.24, 99.68, 99.73 respectively), with a total of 712,875 SNPs homogenously distributed along the genome. We used pedstat from Merlin package to detect the 108 kb deleted region. Whole genome sequencing was performed in the patient and his parents using an Illumina NovaSeq6000 platform. DNA was prepared using Illumina TruSeq DNA PCR-Free library preparation kits, according to the manufacturer’s instructions. An average sequencing depth of 30x was obtained for each sample. Alignment was done using BWA and samtools packages with GRCh37+decoy of human genome referency. BAM file was analysed and pairbase limits of the deletion was determined using Interactive Genome Viewer (IGV).

The 108 kb deletion boundaries were confirmed by Sanger sequencing using primers flanking the deletion (forward: AACAAACCTCCAAGCCACAG; reverse: TTAGGGCTTTCCTCAGCACA), amplifying a 594 bp PCR product in case of deleted allele. PCR amplification of family members using these primers also confirmed the paternal transmission of the deletion in the family (father and child: amplified; mother: not amplified).

### Stem cell culture

The hESC culture and pancreatic differentiation were authorized by the Robert Koch Institute within the “79. und 193. Genehmigung nach dem Stammzellgesetz, AZ3.04.02/0084 und AZ 3.04.02/0190”. hESC line HUES8 (Harvard University, RRID: CVCL_B207) was cultured in Matrigel-coated (hESC-qualified, Corning, #354277) plates in mTeSR Plus medium (STEMCELL technologies, #100-0276) supplemented with 1% penicillin-streptomycin (P/S; Sigma-Aldrich, P4333) at 37°C, 5% CO_2_, 5% O_2_, and humidified atmosphere. The medium was changed every other day. Cells were split at 80-95% confluence with TrypLE Select (Gibco, #A1217701) and seeded on Matrigel-coated plates in mTeSR Plus with P/S and 10 µM ROCK inhibitor Y-27632 (STEMCELL technologies, #72307) as previously described^33^.

### CRISPR-Cas9-based genome editing in hESC

In this study, the heterozygous and homozygous *ONECUT1* hESC (HUES8) knockout lines from Philippi et al. were used^7^. The patient-specific 108 kb deletion was induced in both HUES8 wildtype and heterozygous *ONECUT1* knockout lines using CRISPR-Cas9 gene editing technology. Two crRNAs were designed using the open-access online designing tool CRISPOR^34^ (http://crispor.tefor.net) (guide 1: TATCTCGGGTACCGTCATCC; guide 2: GGATAGGCCATAGTCCATCA; IDT) to create two double strand breaks flanking the region of the Pat1-deletion, resulting in a complete knockout of 108 kb. Modified single guide RNAs (sgRNAs) combining tracrRNA and the respective crRNA were obtained from Synthego. Both sgRNAs and Alt-R^TM^ S.p. HiFi Cas9 Nucelase C3 (IDT, 1081060) were introduced into HUES8 or heterozygous *ONECUT1* cell lines via nucleofection using a 4D Nucleofector and the P3 Primary Cell 4D-Nucleofector X Kit (Lonza, V4XP-3032) following the manufacturer’s instructions. In brief, 200,000 cells were harvested at 60-70% confluency, and resuspended in 16.4 µl P3 solution and 3.6 µl P3 supplement. An RNP complex, consisting of 12 pmol Cas9 and 72 pmol of each sgRNA, was formed during a 10-minutes incubation at room temperature. RNP complex was added to the cell suspension and transferred to one well of a 16-well Nucleocuvette Strip and pulsed with the nucleofection code CB-150. 80 µl mTeSR Plus medium with 10 µM ROCK inhibitor was added and transfected cells were incubated for 5 minutes at 37°C and subsequently seeded on a Matrigel-coated 48-wells. After 48 hours cultivation at 37°C, cells were seeded at low density to obtain single cell-derived colonies within 8-10 days. The mTeSR Plus medium was supplemented with 1x CloneR (STEMCELL technologies, #0588) to enhance cell survival. Colonies were manually isolated for expansion and genotyping of the gene-edited clones.

### PCR for clone selection

Genotyping of the transfected single cell-derived clones was performed using Phire Hot Start II DNA Polymerase without prior DNA isolation. Each 20 µl reaction consisted of 10 µl 2x Phire Tissue Direct PCR Master Mix (Thermo Fisher, F170L), 125 nM forward primer, 125 nM reverse primer, 1 µl of the respective cell suspension and nuclease-free H_2_O. Three primer combinations were used for screening: (1) Primer 1 (CTCCATTGGGGCCTACAAGT, Biomers) and primer 2 (ACTCACAGGCCATCTTGCTC, Biomers) flanking the 5’-edited region of the deletion, generating a PCR product of 849 bp only if the cell line is either wildtype or heterozygous. (2) Primer 3 (ACTTTCTGTTTTAGGACACCTCT, Biomers) and primer 4 (TCCCCCTCATTTCTTGTGTCC, Biomers) flanking the 3’-edited region, generating a 1180 bp product if the clonal cell line is either wildtype or heterozygous. (3) Primer 1 and primer 4, producing a 1184 bp product only if the 108 kb deletion was successfully introduced, whether heterozygous or homozygous. The respective genotypes were verified by Sanger sequencing (Eurofins Genomics).

### Pancreas differentiation of hESCs

#### Differentiation to pancreatic progenitors

Differentiation of hESCs to pancreatic progenitors was performed as previously reported^33^. In brief, cells were seeded on growth-factor reduced (GFR) Matrigel-coated plates (Cultrex UltiMatrix, bio-techne, BME001 or Matrigel, Corning, 354230) in mTeSR Plus medium with 10 µM ROCK inhibitor. Differentiation was initiated after 24 hours at a confluency of 70-95% by adding day 0 medium. The differentiating cells were maintained at 37°C, with 5% CO_2_, and the differentiation medium containing stage-specific cytokines and growth factors was replenished daily. Cells were harvested and analyzed at distinct stages: definitive endoderm (DE; day 3), gut tube endoderm (GTE; day 6), pancreatic endoderm (PE; day 9) and pancreatic progenitor (PP; day 13).

For flow cytometry analysis of NKX6.1 and PDX1 expression on day 13, the protein kinase C activator indolacam V was excluded from the differentiation medium. This adjustment was made to reduce differentiation efficiency, thereby enhancing the detection of genotype-specific differences.

#### Differentiation to endocrine progenitors

Following the differentiation of hESC into pancreatic progenitors as outlined above, the cells were subsequently guided towards pancreatic endocrine progenitors using a previously established monolayer culture method^33,35^. Briefly, at day 13 of differentiation, medium change was performed with stage 5 monolayer medium supplemented with 1 µM Latrunculin A (Biomol, #Cay10010630). After 24 hours, medium was replaced with stage 5 monolayer medium deprived of Latruculin A. Daily medium changes were maintained until reaching the endocrine progenitor stage (day 20), when cells were harvested for downstream analysis.

#### Differentiation to stem cell-derived islets (SC-islets)

To generate mature and functional SC-islets, PPs underwent further differentiation as previously described^28,36^. Initially, PPs were dissociated with TrypLE for 9 minutes at 37°C. The reaction was stopped with DMEM, followed by centrifugation at 180 x g for 5 minutes. After removing the supernatant, the cell pellet was resuspended in stage 5 suspension medium supplemented with 10 µM ROCK inhibitor. Subsequently, cells were counted and 5 ml cell suspension of 1.2×10^6^ cells/ml (6×10^6^ cells per well) was seeded into one well of an AggreWell 400 microwell culture plate (STEMCELL technologies, #34460) pretreated with Anti-Adherence Rinsing Solution (STEMCELL technologies, #07010) according to the manufacturer’s recommendations. AggreWell plates were then centrifuged at 100 x g for 3 minutes and subsequently incubated at 37°C to facilitate sphere formation. After 24 hours, 2 ml of stage 5 medium with 10 µM ROCK inhibitor was added carefully. The following 2 days, 4 ml medium change was performed. Afterwards, spheres of each AggreWell were transferred to individual wells of a 6-well suspension plate, and medium was switched to 4 ml of stage 6 suspension medium. Spheres were then cultured on an orbital shaker at 100 rpm, with daily medium changes of stage 6 suspension medium for a period of 7 days. Subsequently, SC-islets were cultured for over 3 weeks in stage 7 medium to promote maturation, with medium changes performed every other day.

### Insulin secretion

Insulin secretion assay was performed in a 24-well plate, utilizing approximately 20 to 30 carefully selected spheres. Each experimental condition comprised 3 to 6 replicates from 2 or 3 independent differentiation processes. Spheres were rinsed twice with freshly prepared KRB buffer^37^ and incubated in 200 µl low glucose (LG) condition (KRB + 1 mM glucose) for 90 minutes at 37°C on an orbital shaker. Supernatant was discarded, spheres were washed twice with KRB buffer and sequentially incubated for 30 minutes in the following conditions: LG condition; high glucose (HG) condition (KRB + 20 mM glucose); HG + 50 ng/ml exendin-4 (MedChemExpress, HY-13443); LG condition, LG + 30 mM KCl. For stimulation with tolbutamide, the spheres were equilibrated for 90 minutes in LG condition, following 30 minutes stimulation with LG condition and 30 minutes LG + 400 µM tolbutamide (MedChemExpress, HY-B0401). Subsequently, the supernatants were collected, and the secreted insulin content was quantified using an insulin enzyme-linked immunosorbent assay (ELISA; Mercodia, #10-1113-10) according to the manufacturer’s protocol. Samples were appropriately diluted 1:10 (KCl condition) or 1:5 (all other conditions). The spheres were dissociated and counted using a CASY cell counter (OLS) to normalize the secreted insulin levels to the respective cell numbers.

### Flow cytometry

#### Surface marker staining

Cells were harvested, washed with PBS, followed by incubation in 300 µl FC buffer containing 10% Panexin (PAN-biotech, #P04-96950) in PBS for 30 minutes on ice. Next, cells were centrifuged at 1000 x g, resuspended in 50 µl primary antibody diluted in FC buffer, and incubated overnight at 4°C. Afterward, the cell pellet was washed twice with FC buffer, then resuspended in 50 µl secondary antibody (Donkey-anti-mouse, Alexa Fluor 488-, 568-, 647-conjugated, Thermo Fisher) diluted 1:500 in FC buffer, and incubated for 90 minutes on ice. If the primary antibody was directly coupled to a fluorochrome, this step was omitted. The cells were washed twice with FC buffer, resuspended in 300 µl FC buffer and measured using Attune NxT flow cytometer (Thermo Fisher).

The following primary antibodies were used: anti-SSEA4 1:10 (PE-conjugated; BD biosciences; #560128) and anti-TRA-1-60 1:10 (FITC-conjugated; BD biosciences; #560380) for hESC stage, and anti-GP2 1:5000 (MBL international; D277-3) for PP stage.

#### Intracellular marker staining

Intracellular flow cytometry staining was performed as previously described^33,38^. The following primary antibodies were used: anti-NKX6.1 (1µg/ml, DSHB, F55A12), anti-PDX1 (1:250, R&D, AF2419), anti-SOX17 (1:500, R&D, #AF1924), anti-GATA6 (1:500, Cell Signaling, #5851), anti-GCG (1:500, Sigma, #G2654), anti-CPEP (1:100, Cell Signaling, #4593), anti-INS-647 (1:80, AF647-conjugated, Cell Signaling, #9008). Alexa Fluor 488-, 568-, or 647-conjugated secondary antibodies (Thermo Fisher) were used in 1:500 dilution.

For endocrine progenitor staining, cells were additionally incubated in NKX6.1-647 conjugated antibody (1:150) for additional 90 minutes on ice. Conjugation of NKX6.1 antibody was performed with FlexAble CoraLite® Plus 647 Antibody Labeling Kit (Proteintech, #KFA003) according to the manufacturer’s protocol. Analysis of FC measurements was performed with FlowJo software (V10.10.0).

### Immunofluorescence staining using Cytospin

Cells were harvested and dissociated with TrypLE, fixed in 4% PFA / 100 mM sucrose for 25 minutes on ice and stored in PBS until further processing. 80 µl cell suspension was centrifuged onto SuperFrost Plus Glass slides (VWR) with a Cytospin Centrifuge (Thermo Fisher) for 3 minutes at 600 rcf. Cells were circled with a hydrophobic pen and subjected to blocking and permeabilization in blocking buffer (5% normal donkey serum (NDS), 0.1% Triton-X, PBS) for 30 minutes at room temperature. After removal of the liquid, cells were incubated in primary antibody diluted in blocking solution at 4°C overnight. Cells were washed three times with wash solution (2% NDS, 0.1% Triton-X, PBS) for 5 minutes and incubated subsequently in secondary antibodies (Alexa Fluor 488- or 568-conjugated secondary antibodies; Thermo Fisher) diluted 1:500 in blocking buffer along with 500 ng/ml DAPI and incubated for 90 minutes at room temperature. Cells were washed three times with wash buffer for 5 min. For GP2 (1:1000) surface staining, blocking and wash buffer was replaced by 10% Panexin in PBS, while the staining procedure remained unchanged. Finally, cells were mounted with Fluoromount-G (SouthernBiotech) and imaged using a Zeiss Axioscope2 microscope (Carl Zeiss).

### Western blot

Western blot for ONECUT1 and β-Actin was performed on PP samples as previously described^7^. In brief, 40 µg protein was separated on a 10% SDS-PAGE and transferred on a PVDF membrane using a semidry transfer system (Bio-Rad). The following antibodies were used: anti-ONECUT1 (1:2000, Santa Cruz; H1-100-X, #sc-13050), anti-β-Actin (1:5000, Sigma, #A5316), anti-mouse-HRP and anti-rabbit-horseradish peroxidase (HRP; GE healthcare; #NA931, #NA934). HRP detection was performed using SuperSignal West Duro Kit (Thermo Fisher Scientific) and Chemiluminescence Imaging Fusion Solo X system (VILBER Lourmat). Western blot bands were quantified using Fiji (ImageJ) software^39^.

### RNA isolation, reverse transcription, qPCR

RNA was isolated from PP cells using GeneJET RNA Purification Kit (Thermo Fisher Scientific) following the manufacturer’s protocol. Isolated RNA was either used for bulk RNA sequencing or for reverse transcription and qPCR. Reverse transcription was performed with Scriptase RT cDNA Synthesis kit (Genaxxon). qPCR was done with GreenMasterMix No ROX (Genaxxon) with Rotor-Gene Q (Qiagen).

The following primers were used: lncRNA-AC forward (TCGTGGTGGAGGTTTTGAGA, Biomers), lncRNA-AC reverse (TCTGCCCTTCTGCTGTCATT, Biomers), Hs_HMBS_1_SG (QT00014462, Qiagen), Hs_ONECUT1_1_SG (QT00219352, Qiagen).

### RNA fractionation for subcellular localization analysis

Cells were harvested for RNA extraction. Half of the sample was further processed for total RNA isolation as described before. RNA isolation of the remaining cell portion was conducted using the RNA Subcellular Isolation Kit (ActiveMotif, #25501) according to the protocol provided by the supplier, which facilitates the separation of nuclear and cytosolic RNA. Equal amounts of cytosolic, nuclear, and total RNA were used for reverse transcription and qPCR as described. Ct values of cytosolic and nuclear fractions were normalized to the lncRNA-AC Ct value of the respective total RNA sample.

### Immunofluorescence staining on paraffin sections

SC-islets were fixed in 4% PFA / 100 mM sucrose overnight at 4°C. Spheres were pre-embedded in 2% agarose and underwent standard histological procedures. This included automated serial dehydration, followed by paraffin embedding and sectioning (4 µm). Immunofluorescence staining was performed as previously described^38^. The following primary antibodies were applied: anti-INS (1:5000, abcam, #ab181547), anti-GCG (1:1000, Sigma, #G2654), anti-CHGA (1:200, DAKO, #M0869), anti-SOX9 (1:500, Millipore, #AB5535), anti-VIM (1:500, Cell Signaling, #5741S), anti-PPY (1:500, proteintech, #15493-1-AP), anti-SST (1:200, proteintech, #24496-1-AP). Alexa Fluor secondary antibodies (Thermo Fisher) were used. Slides were imaged with Zeiss Axioscope2 microscope (Carl Zeiss). Image adjustments including cropping and brightness/ contrast enhancements were carried out using Fiji (ImageJ) software^39^. Quantification was performed utilizing QuPath (V0.5.0)^40^.

### Transmission Electron Microscopy

SC-islets were fixed overnight at 4°C in 4% PFA / 100 mM sucrose and stored in PBS. Sample preparation for transmission electron microscopy was carried out by the Central Facility for Electron Microscopy at Ulm University following the protocol as previously reported^41^. The islets were fixed in 0.1 M phosphate buffer (pH 7.3) with 2.5% glutaraldehyde and 1% sucrose. The samples were post-fixed in 2% aqueous OsO_4_ and dehydrated in a graded series of isopropanol. The samples were then block-stained with 1% uranyl acetate in ethanol and embedded in Epon. Ultrathin sections (80 nm) were cut with an ultramicrotome equipped with a diamond knife, transferred to copper TEM grids and post-stained with 0.3% lead citrate for one minute. Imaging was performed with a JEOL TEM 1400.

### RNA-seq

For bulk-sequencing of poly(A)-RNA, library preparation was conducted as described previously^42^. Barcoded cDNA of each sample was generated with a Maxima RT polymerase (Thermo Fisher) using oligo-dT primers containing barcodes, unique molecular identifiers (UMIs) and an adapter. 5’-ends of the cDNAs were extended by a template switch oligo (TSO) and full-length cDNA was amplified with primers binding to the TSO-site and the adapter. NEB UltraII FS kit was used to fragment cDNA. After end repair and A-tailing, a TruSeq adapter was ligated, and 3’-end-fragments were finally amplified using primers using Illumina P5 and P7 overhangs. Sequencing of the library was performed on a NextSeq 500 (Illumina) with 61 cycles for the cDNA in read1 and 19 cycles for the barcodes and UMIs in read2. Data processing utilized the published Drop-seq pipeline (v1.0) to generate sample- and gene-wise UMI tables^43^. Reference genome (GRCh38) was used for alignment. Transcript and gene definitions were used according to GENCODE v38.

DESeq2 version 1.38.3 was utilized to normalize raw counts, and a filtering step was applied to retain genes with a minimum count of 10 across at least 3 samples. Subsequently, a variance stabilizing transformation was performed for downstream analyses, including principal component analysis (PCA). Differential expression analysis between genotypes was conducted using the “result” function, employing a significance threshold of p < 0.05.

### GSEA

For gene set enrichment analysis, the GSEA desktop software version 4.3.3 was employed, utilizing the GSEAPreranked tool. In this analysis, differentially expressed genes (DEGs) were ranked based on the negative logarithm (base 10) of their p-values multiplied by the sign of the corresponding log2 fold change. GSEA was performed using specific endocrine lineage gene sets^31,32^.

### GapmeR-based lncRNA knockdown

GapmeR sequences were designed with the Antisense LNA GapmeR Custom Builder online tool (Qiagen). Two GapmeRs from top-ranked sequences were selected.

LncRNA-AC knockdown was performed in HUES8 wildtype cells that were differentiated into endocrine progenitors in 48-well format as described above. Lipofection was performed on days 9, 11, 13, 15, 17, and 19 of differentiation, with cell harvesting performed on day 20 for downstream analysis. The transfection protocol involved mixing 25 µl DMEM and 0.5 µl Lipofectamine2000 (Thermo Fisher) added to a second tube containing 25 µl DMEM with 45 pmol GapmeR-1 (AC1), GapmeR-2 (AC2) or negative control GapmeR. Transfection mix was incubated for 15 minutes at room temperature. Meanwhile differentiation medium was changed. Lipofection mix was added dropwise to the wells. Subsequent medium changes were performed not earlier than 24 hours after lipofection. A FAM-labeled negative control LNA GapmeR (Qiagen) was used to monitor transfection efficiency and to exclude general effects based on LNA GapmeR transfection.

### Luciferase assay

Luciferase assay was performed as previously described^7^. Enhancer sequences, containing an EcoRV restriction sites at both ends, were synthesized and inserted into a pUC vector backbone by Azenta Life Sciences. These sequences were cloned into the pGL3-enhancer vector (Promega, GenBank Accession Number U47297) after SmaI digestion. Due to blunt end ligation, the insertion of the sequence could occur in either the minus or plus orientation. Confirmation of the correct sequence and orientation was achieved through a control digest. For reporter gene assays HeLa cells (2×10^5^ cells) were seeded in a 48-well plate 16 hours prior to transfection with Lipofectamine 2000. Transfection included 250 ng of the enhancer plasmid along with or without a respective expression plasmid (ONECUT1^7^) at varying concentrations (50 ng or 100 ng). Cells were lysed after 24 h, and luciferase activity was measured in 10 µl cleared lysate using a Centro LB960 luminometer (Berthold Technology) and the luciferase assays system (Promega).

### Hi-C

Here, we used the Hi-C data from the regulation of cohesin loop extrusion, which underlies changes in enhancer-promoter interactions during pancreatic cell differentiation^44^. Hi-C files were obtained from GSE210524 (https://www.ncbi.nlm.nih.gov/geo/query/acc.cgi?acc=GSE210524). Subsequently, we converted the coordinates from hg38 to hg19 to align with our previous sequencing data^12^ using Hiclift^45^. We detected structural differences between stem cell (H9 cell line), DE and PP using CHESS^46^. Before, we normalized the data with Knight–Ruiz matrix balancing^47^. CHESS was applied to 5-kb resolution Hi-C matrices using windows of 1 mb and a step size of 1 kb to produce similarity scores for pairwise Hi-C comparisons between ESC and the differentiation stages PP and DE.

### Nanopore-Sequencing

For methylation analysis and variant phasing, nanopore sequencing with adaptive sampling and 5hmC and 5mC methylation calling was conducted. For this purpose, HMW (high molecular weight) DNA was extracted using the Promega Wizard Genomic DNA Purification Kit for cultured cells (Promega, #A1120), according to the manufactureŕs manual. The DNA was dissolved overnight at 4°C and quantified by using the Quant-IT^TM^ dsDNA Assay kit (ThermoFisher Scientific, #Q33232). Approximately 2.5 - 4.5 µg of the extracted DNA was sheared using g-TUBEs (Covaris, #520079) and used as input for nanopore library preparation with the ligation sequencing DNA V14 kit (ONT, SQK-LSK114) according to the manufactureŕs protocol. Each library was loaded on a MinION R10.4.1 flow cell (ONT, FLO-MIN114) and sequenced with a MinION Mk1B device for up to 72 h. Throughout the sequencing process, the flow cells were reloaded 1-2 times after a nuclease flush (ONT, EXP-WSH004) with the same library according to the manufactureŕs instructions to enhance sequencing yield. For adaptive sampling a more than 800 kb region (chr15.hg19: 52999842_53878937) with the “fast modified basecalling for 5hmC and 5mC in CG context” basecall model of the MinKNOW software version 23.07.12 and integrated Guppy version 7.1.4 was set.

### ONT Analyse phasing and methylation

The WT HUES8, WT PP, ONECUT1 -/+ Pat1-del -/+ cis (clone 1), and trans (clone 3) cell lines were subjected to analysis using unaligned BAM files. The Epi2Me human variation workflow (version 1.9.0, available at https://github.com/epi2me-labs/wf-human-variation) was executed to generate DNA methylation data, structural variant data, and phased aligned BAM files (based on the GRCh38 reference genome) for the genomic region spanning coordinates chr15.hg38:52707645_53586740. A minimum coverage threshold of 1 was applied. Additionally, the BAM files underwent manual inspection using the Integrative Genomic Viewer (IGV, version 2.17.0).

### Statistics

Statistical tests were performed with GraphPad prism software v10.0. For comparison of two groups unpaired t-test (two-tailed) was performed. Analysis of several groups was performed with ordinary one-way ANOVA. Bar graphs depict mean ± standard error of the mean (SEM). Significance was displayed as follows: p < 0.05: *, p < 0.01: **, p < 0.001: ***, p < 0.0001: ****.

### Data availablility

Sequencing data that have been generated for this study will be deposited at the Gene Expression Omnibus (GEO) before publication. Hi-C data that were used for reanalysis were obtained from GSE210524. ChIPSeq, histone marks abd ATAC peaks from wildtype and ONECUT1 deleted hESCs from gut tube, pancreatic endoderm and pancreatic progenitor stage were obtained from GSE131817.

### Code availablility

Programs used for transcriptome analysis, Hi-C data reanalysis and ONT analyse phasing and methylation are described in Methods section in more detail.

---

**Extended Data Fig. 1:**
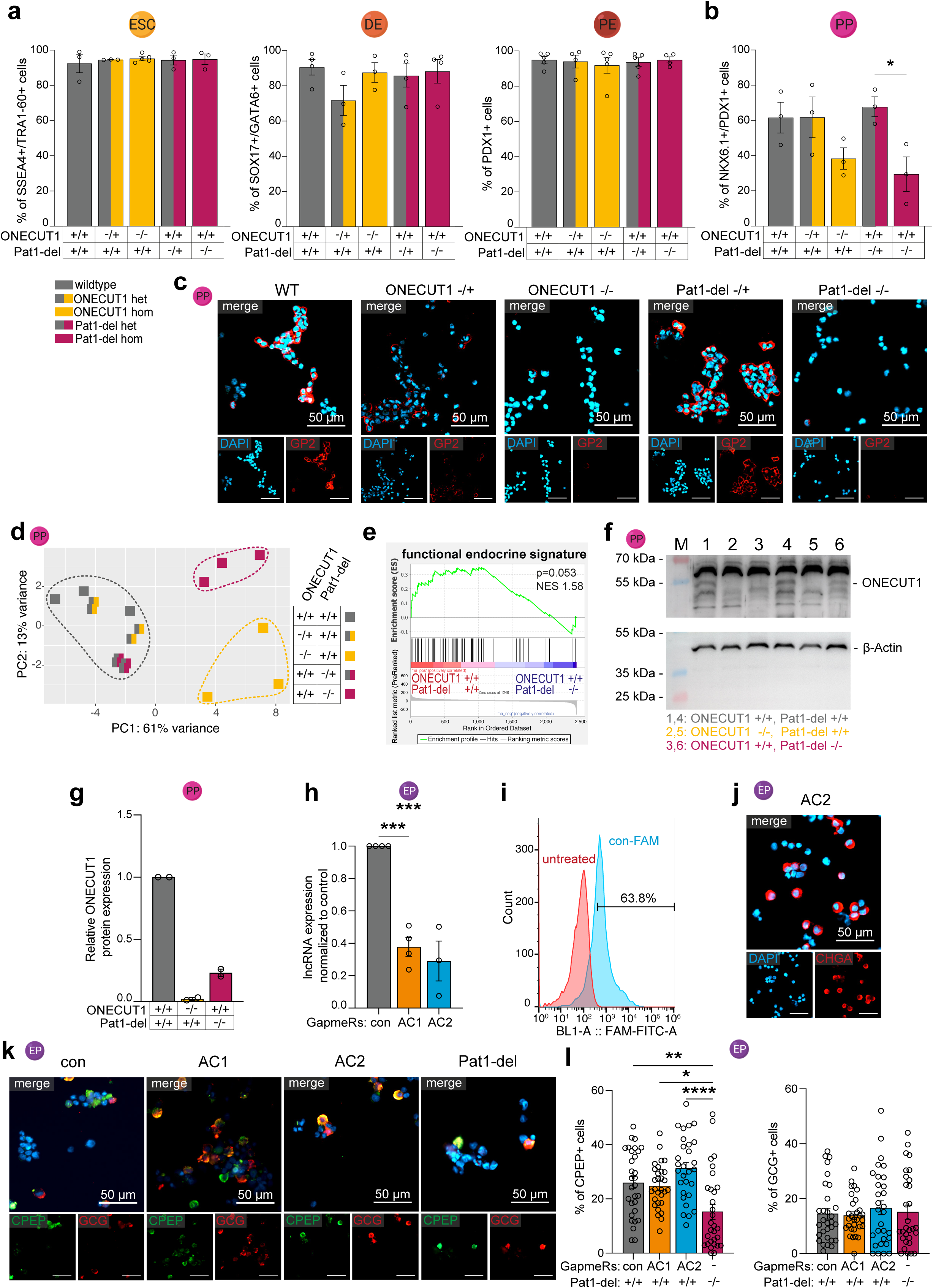
**(a)** Flow cytometry of stem cell marker SSEA4 and TRA1-60 of indicated genotypes (n=3-4). Pancreatic differentiation was monitored at definitive endoderm (DE) for SOX17 and GATA6 (n=3-4), at pancreatic endoderm (PE) for PDX1 (n=4-5) and at (**b**) pancreatic progenitor stage for NKX6.1 and PDX1 (n=3; indolactam V omitted from culture medium). (**c**) Representative immunofluorescence (IF) images of glycoprotein 2 (GP2) at PP stage. (**d**) Principal component analysis (PCA) of whole transcriptomes of indicated genotypes. (**e**) GSEA proves loss of a functional endocrine gene set in Pat1-del -/- PPs^32^. (**f**) Western blot analysis for ONECUT1 in PP samples of wildtype and ONECUT1 homozygous or Pat1-del lines, (**g**) including the respective quantification (n=2). (**h**) qPCR showing successful knockdown by AC1 and AC2 GapmeR, normalized to GapmeR negative control (n=4). (**i**) Flow cytometry histogram confirming successful transfection of differentiating cells with GapmeRs, indicated by FAM-fluorophore detection (coupled to negative control). (**j**) Chromogranin A (CHGA) IF staining in lncRNA-AC knockdown cells with GapmeR AC2. (**k,l**) Representative IF images of glucagon (GCG) and c-peptide (CPEP) and the respective quantification.

**Extended Data Fig. 2:**
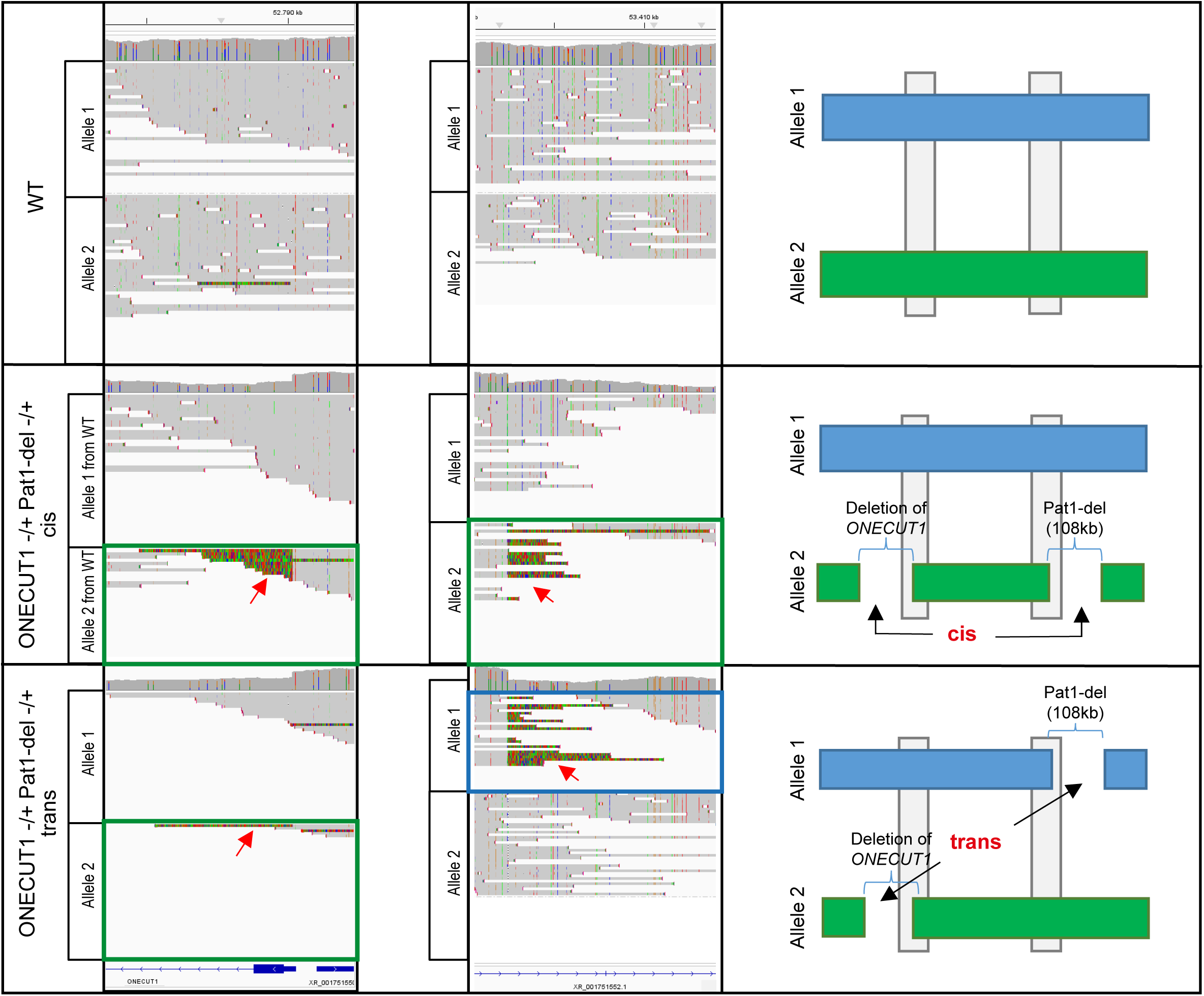
Allelic assignment of deletions introduced by CRISPR-Cas9 into wildtype HUES8 cells. IGV viewer display of ONT sequencing reads with colour labelling of variants according to base composition (green = adenine, blue = cytosine, brown = guanine, red = thymine; left) and schematic illustrations of allele 1 (blue bars) and allele 2 (green bars) for wildtype and ONECUT1 -/+ Pat1-del -/+ in cis (clone 1) and trans (clone 3) (right). The grey boxes represent the areas of the displays in IGV viewer. Top Panel: Discrimination of both alleles of the *ONECUT1* locus (chr15.hg38:52783584_52792381; left) and of the start of the Pat1-deletion (chr15.hg38:53400784_53414213; middle). Middle Panel: Soft clipped reads (coloured) at both ends of the non-deleted part between the *ONECUT1* and the Pat1-deletion shows that the deletions involved are located on both sides of the wildtype allele 2 in ONECUT1 -/+ Pat1-del -/+ cis (clone 1), i.e. the deletions are on the same allele (allele 2, in cis). Lower Panel: Soft clipped reads (coloured) at both ends of the non-deleted part between the *ONECUT1* and the Pat1-deletion shows that the deletions involved wildtype allele 2 (left) and wildtype allele 1 (middle) in ONECUT1 -/+ Pat1-del -/+ trans (clone 2), i.e. the deletion are on two different alleles (in trans).

**Extended Data Fig. 3:**
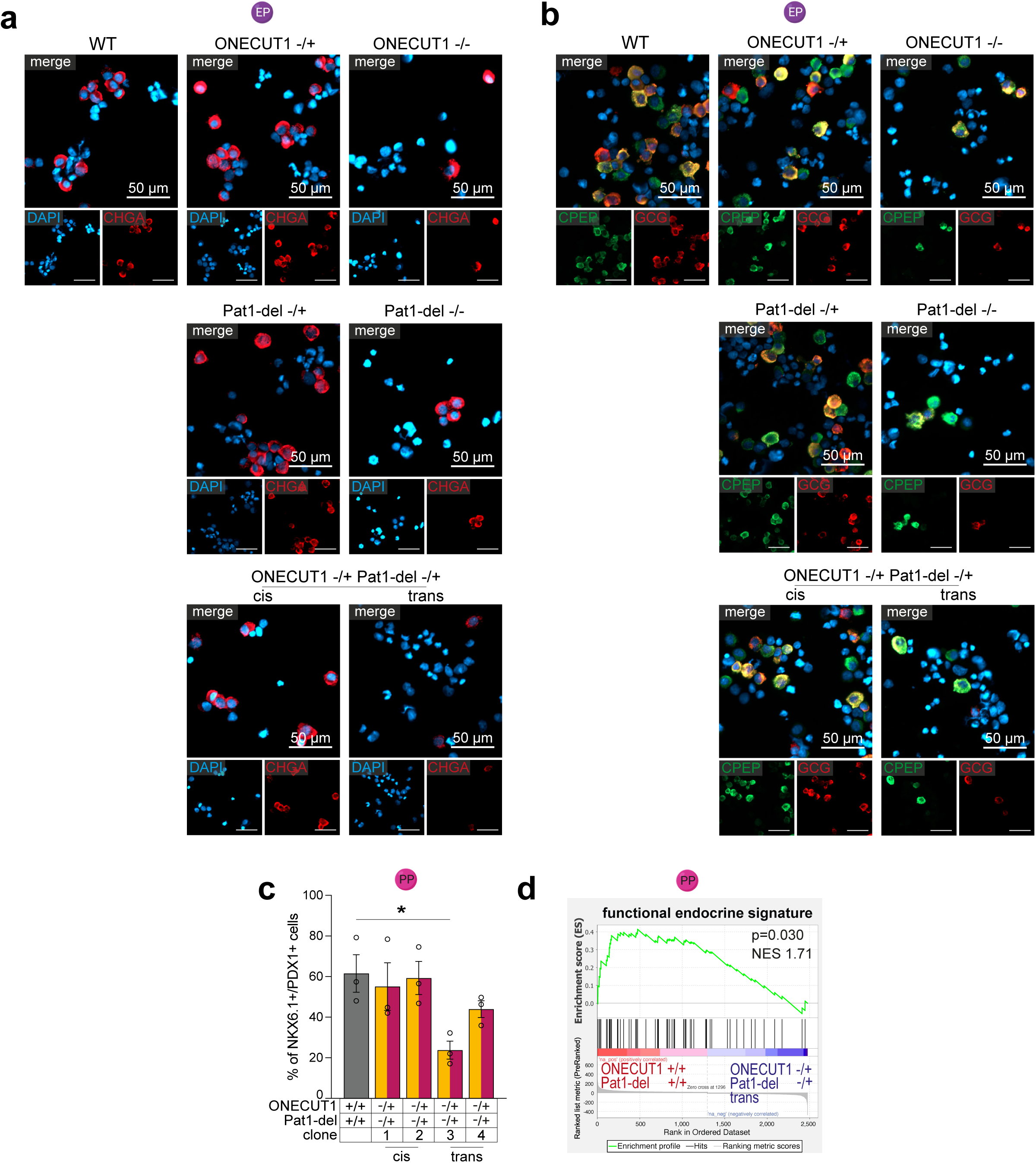
(**a, b**) Immunofluorescence staining of endocrine progenitors (EP) of indicated genotypes with (**a**) chromogranin A (CHGA) or (**b**) c-peptide (CPEP) and glucagon (GCG). (**c**) Flow cytometry of pancreatic progenitor (PP) marker NKX6.1 and PDX1 of ONECUT1 -/+ Pat1-del -/+ in trans or cis (differentiation without indolactam V). (**d**) GSEA shows loss of a functional endocrine gene set in Pat1-del -/- PPs^32^.

**Extended Data Fig. 4:**
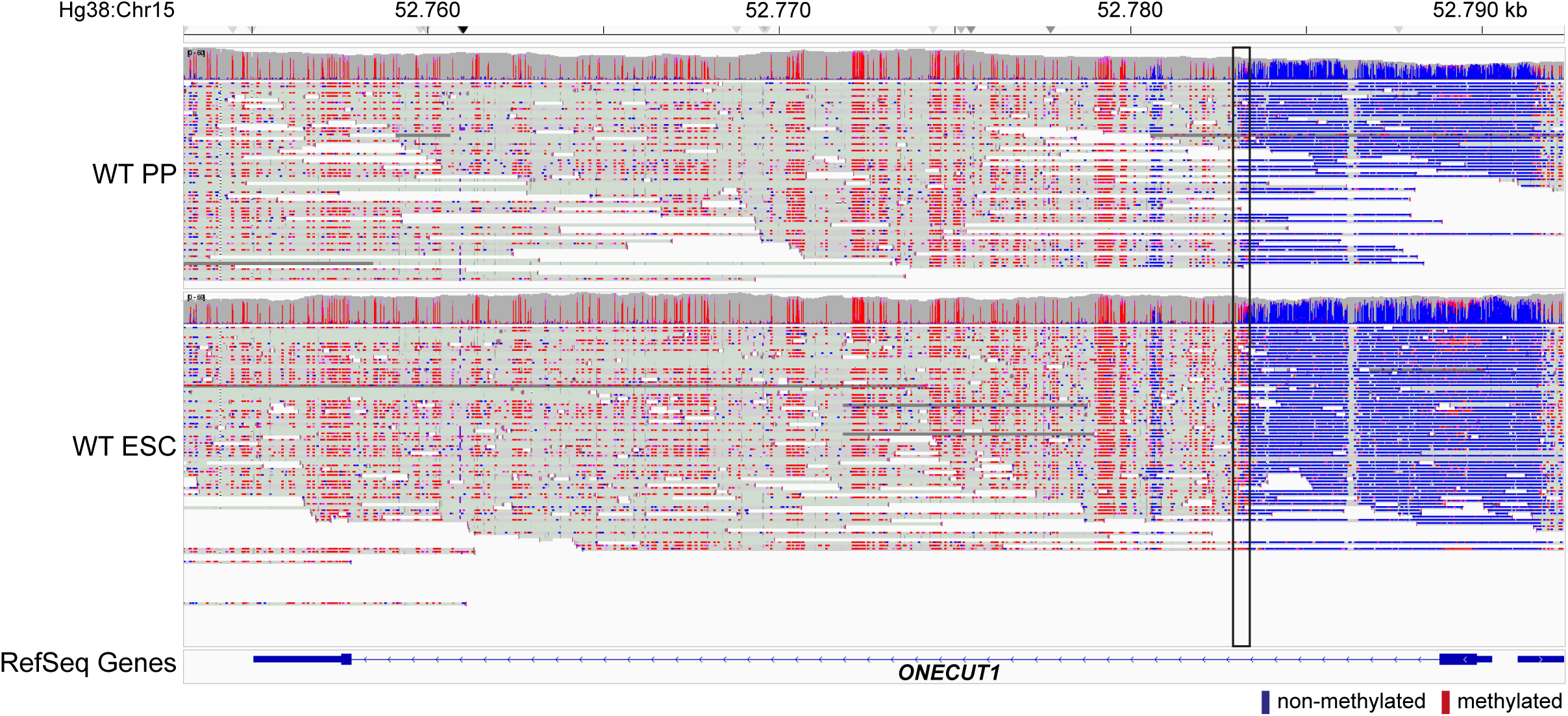
Comparison of the DNA methylation of wildtype HUES8 PP and hESC stage obtained by ONT sequencing and displayed in the IGV viewer. Position chr15.hg38:52753053_52792336 in *ONECUT1* is shown. The black box displays lack of methylation (blue labels) in pancreatic progenitors (PP) as compared to human embryonic stem cells (hESC) which is mostly methylated (red labels) in that region.

**Extended Data Fig. 5:**
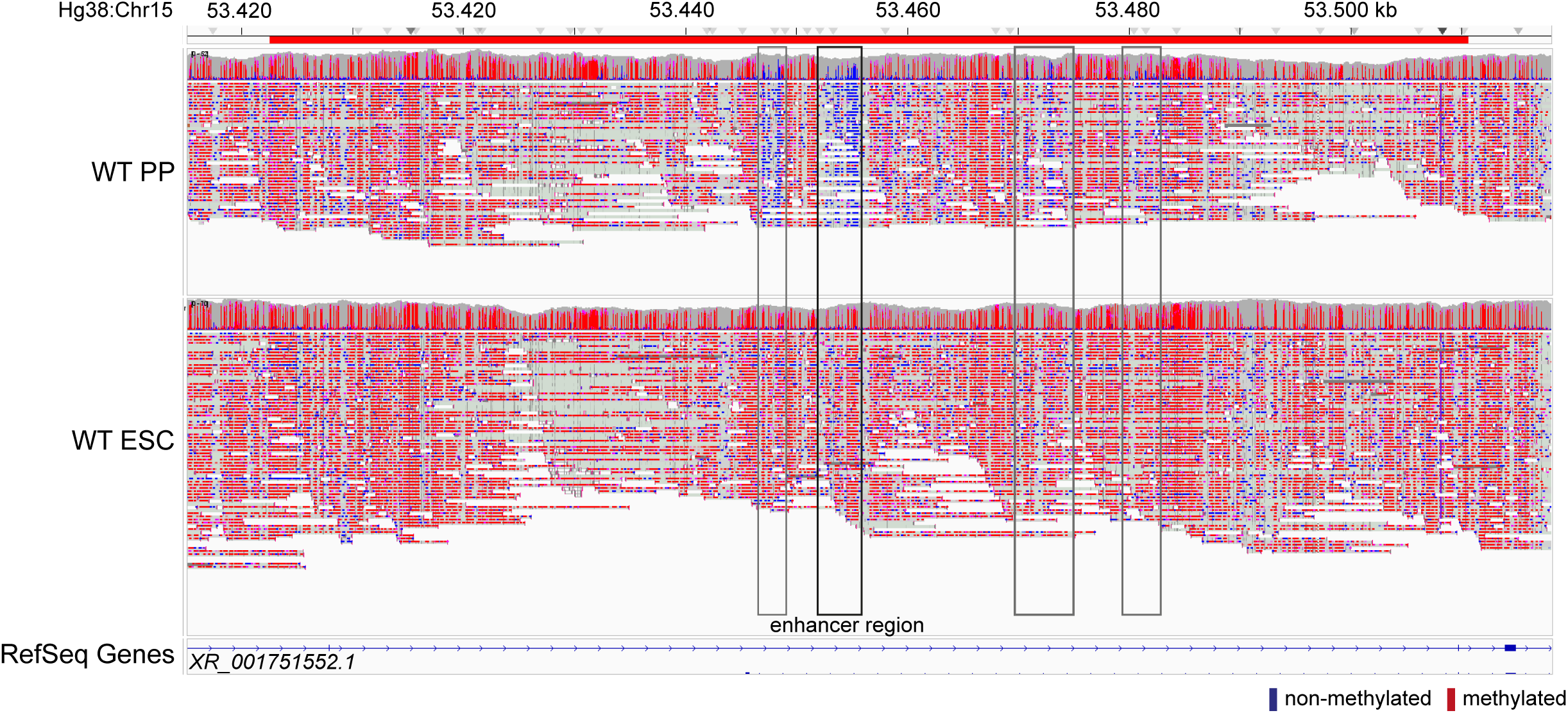
Comparison of the DNA methylation of wildtype HUES8 pancreatic progenitor (PP) and human embryonic stem cell (hESC) stage obtained by ONT sequencing and displayed in the IGV viewer. Position chr15.hg38:53395109_53518044 from the 108 kb sized deleted region is shown (grey box). The proposed enhancer region is highlighted by the black box and displays lack of methylation (blue labels) in PP as compared to hESC which is mostly methylated (red labels) in that region.

**Extended Data Fig. 6:**
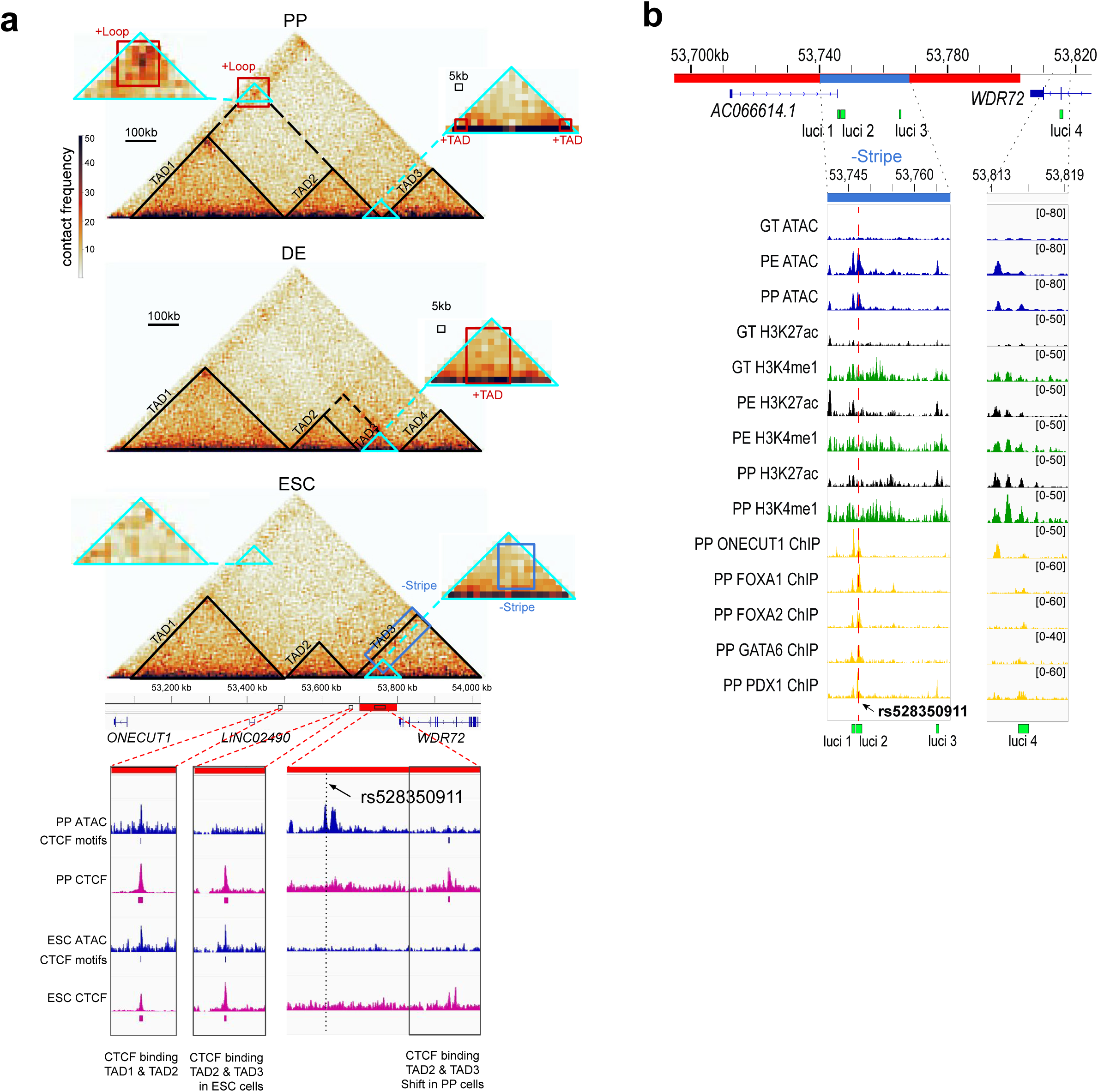
Identification of structural changes between embryonic stem cell stage (ESC; H9) and pancreatic progenitor (PP) / definitive endoderm (DE) cells. Lost and gained structures are highlighted in blue and red squares, respectively. Log2 fold-change maps are depicted below the region of interest with conformational changes. A resolution of 100 kb has been used for feature extraction. The regions of interest were enlarged to a resolution of 5 kb. (**a**) Highly dissimilar regions identified by CHESS between ESC and PP or DE cells. (**b**) IGV plot of stage-specific ATAC-seq, histone marks and Chip-seq of pancreatic transcription factors is shown at selected luci-sites for gut tube (GT), pancreatic endoderm (PE) and PP stage. A SNP rs528350911 is located within the identified enhancer region (luci 2).

**Extended Data Fig. 7:**
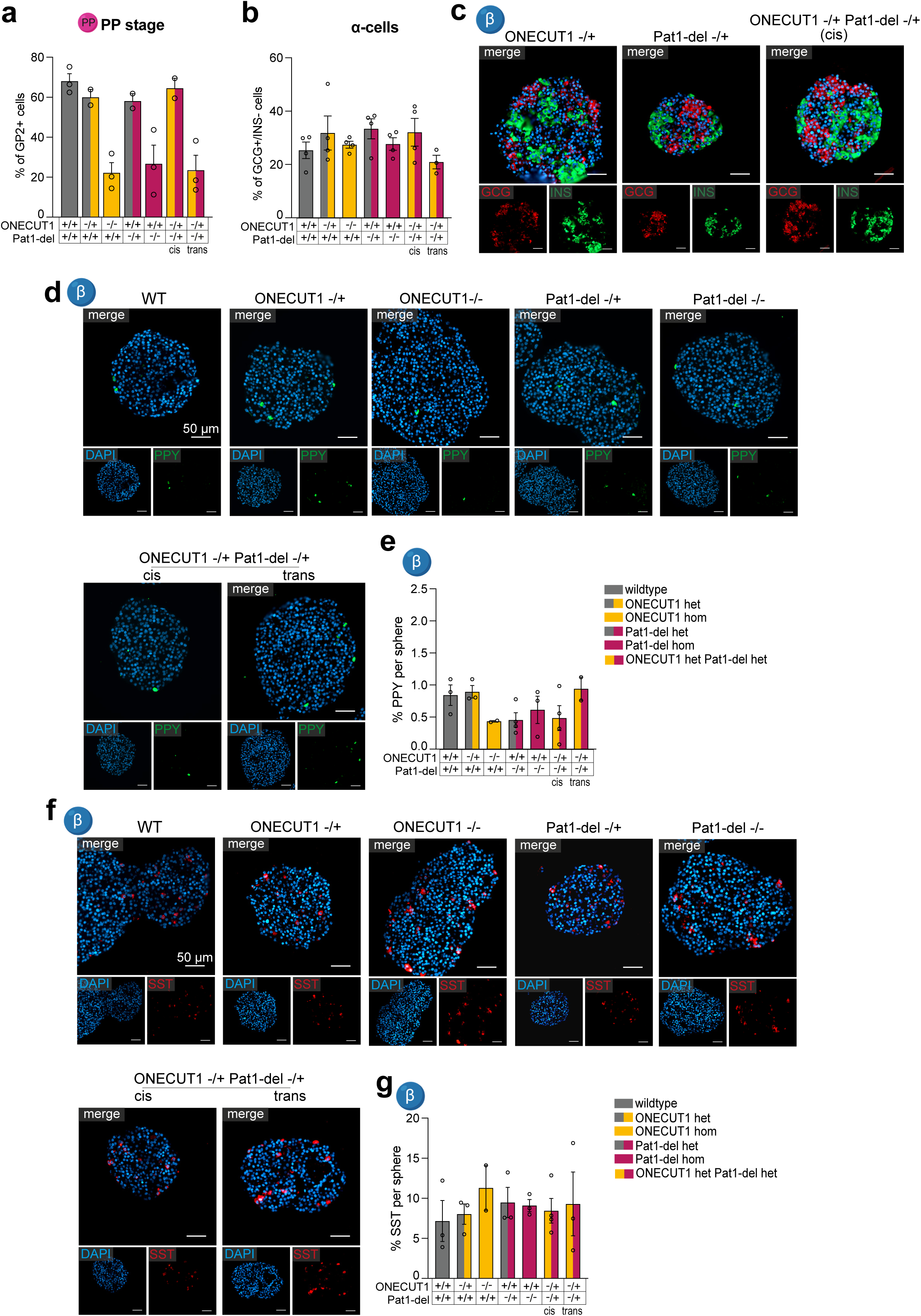
(**a**) Quality control of pancreatic progenitors (PP) used for stem cell-derived islet (SC-islet) differentiations, measuring GP2 positive cells by flow cytometry (n=2-3). (**b**) Percent of alpha-cells in SC-islets of indicated genotype, measured by flow cytometry of GCG+/INS-cells (n=3-4). (**c-g**) Immunofluorescence staining of SC-islets for (**c**) glucagon (GCG) and insulin (INS), (**d**, **e**) pancreatic polypeptide Y (PPY) and (**f**, **g**) somatostatin (SST) and the respective quantifications.

**Extended Data Fig. 8:**
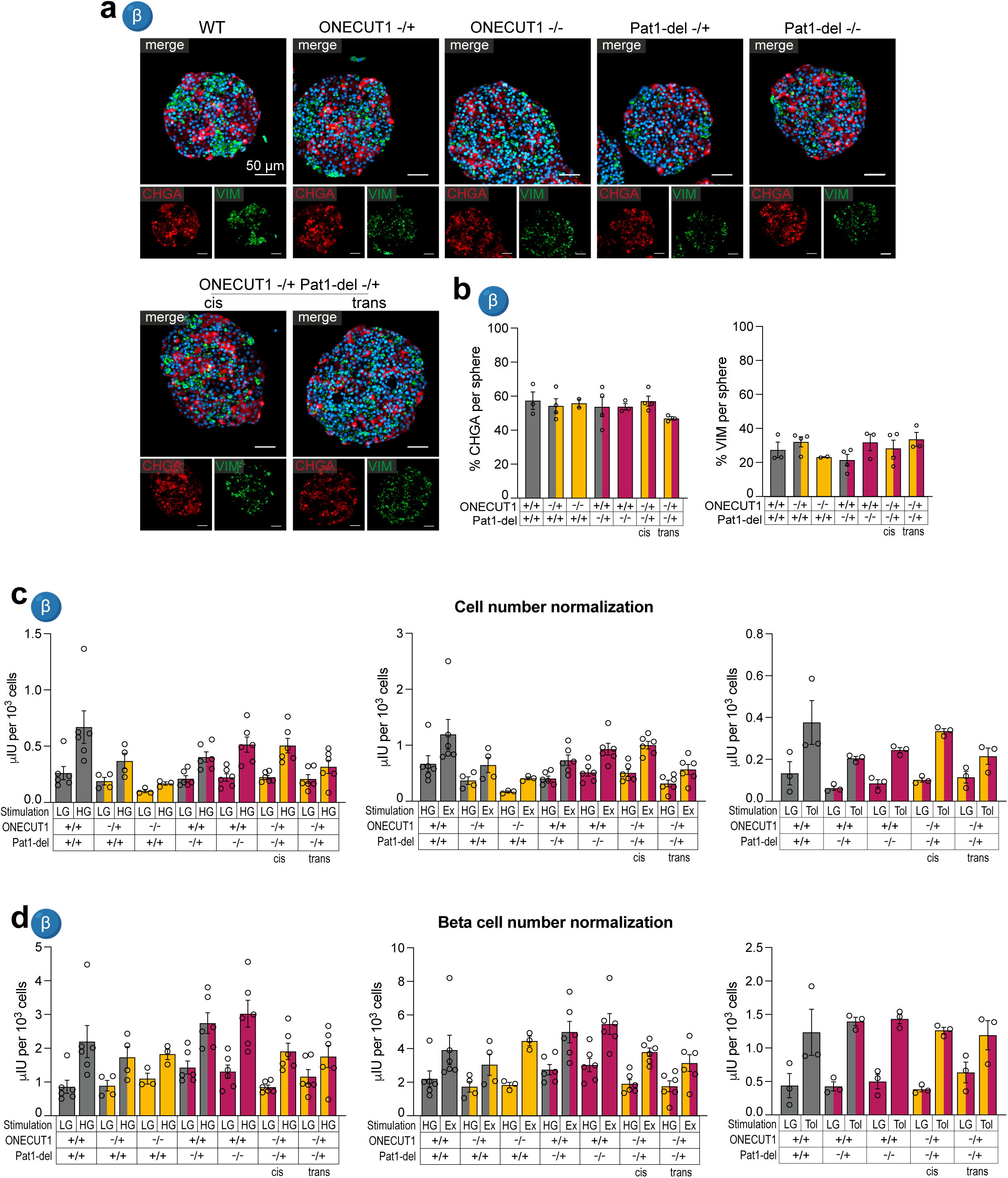
(**a**) Representative immunofluorescence stainings of chromogranin A (CHGA) and vimentin (VIM) and (**b**) the respective quantification. (**c**, **d**) Insulin secretion of stem cell-derived islets (SC-islets) was either serially stimulated by low glucose (LG), high glucose (HG) and HG + exendin-4 (Ex) or LG and LG + tolbutamide (Tol). Secreted insulin is normalized to (**c**) cell number or to (**d**) β-cell number.

## Notes

### Competing Interest Statement

The authors have declared no competing interest.

### Author Declarations

Ethics committee of Montreal (Centre Hospitalier Universitaire Sainte-Justine) gave ethical approval for this work.

